# Transcriptomic profiling during normothermic machine perfusion of human kidneys reveals a pro-inflammatory cellular landscape and gene expression signature associated with prolonged delayed graft function after transplantation

**DOI:** 10.1101/2025.03.28.25324769

**Authors:** HVM Spiers, SA Hosgood, M Larraz, LJKS Stadler, Y Zhai, H Moon, S MacMillan, A Paterson, ML Nicholson, I Mohorianu, V Kosmoliaptsis

## Abstract

Assessment and treatment of severe ischaemia-reperfusion-injury (IRI) remains an unmet challenge in kidney transplantation. Normothermic machine perfusion (NMP) aims to resuscitate organs but also recapitulates IRI *ex situ*. Understanding transcriptional pathways, and associated cellular landscape, driving IRI during NMP could facilitate therapeutic targeting. Using tissue and urine from kidneys undergoing NMP pre-transplantation as part of a randomised controlled trial, we undertook in-depth transcriptomic analyses of kidneys developing prolonged delayed graft function (PDGF; clinical manifestation of severe IRI) versus immediate graft function (IGF). We validated upregulation of previously described pro-inflammatory and immune pathways and identified innate immune system driven processes at the core of the transcriptional signature in PDGF kidneys. Deconvolution using single-cell-RNAseq data showed PDGF kidneys were enriched for pro-inflammatory mononuclear phagocyte, myofibroblast and fibroblast, but depleted of tubuloepithelial, cell signatures. These findings were recapitulated in tissue biopsies from an external NMP kidney cohort with high versus low acute tubular injury, histologically similar to PDGF/IGF kidneys, respectively. Extracellular vesicles released in the urine of PDGF kidneys were enriched in miRNAs functionally annotated to transcriptional processes upregulated in the tissue compartment. Together, our study characterises the transcriptional signature of severe IRI during NMP, highlighting the role of pro-inflammatory innate and pro-fibrotic cells in this process.

## Introduction

Kidney transplantation (KT) represents the optimal treatment for most patients with end stage renal disease. Compared to dialysis, it offers significant benefits to patients in terms of morbidity(1), survival(2,3) and quality of life(1), with further economic advantages for healthcare systems(4). Organ shortage remains a major challenge, with several strategies employed to increase the donor pool, including the use of marginal organs from extended criteria donors (ECD) and donation after circulatory death (DCD) donors. However, marginal organs suffer an enhanced ischaemia-reperfusion injury (IRI), clinically manifesting as delayed graft function(DGF; 5–7). DGF in turn is associated with poor long term outcomes, including worse long term graft function, increased rates of rejection and graft loss(6,7).

Normothermic machine perfusion (NMP) enables organ perfusion with oxygenated nutrient rich solutions in the absence of circulating immune cells and complement. NMP can be used to assess the viability of organs and “recondition” those initially deemed unsuitable for transplant(8). It has been established as feasible and safe in clinical kidney transplantation at the randomised controlled trial level(RCT)(9). NMP recapitulates IRI *ex situ* providing a unique isolated organ platform for the study and manipulation of IRI. In a previous transcriptomic analysis of kidneys during NMP as part of the same RCT, we demonstrated that kidneys going on to develop DGF have a different transcriptional profile compared to those with immediate graft function (IGF)(10). Specifically, during NMP kidneys that went on to develop DGF requiring dialysis in only the first 24 hours post-transplant (commonly for hyperkalaemia) shared transcriptomic profiles with IGF kidneys, whilst those requiring dialysis beyond 24hrs had a distinct inflammatory profile(10).

NMP represents a unique platform for the multiparametric assessment of organs prior to transplantation. In an era of therapeutic delivery during NMP, a validated transcriptomic signature associated with severe IRI (clinically correlated with prolonged DGF) would be valuable in organ assessment and identification of those requiring therapeutic intervention pre-transplant. This signature could also represent a transcriptomic endpoint for therapeutic studies during NMP. Furthermore, granular insights into the mechanisms and cellular landscape of kidneys suffering severe IRI during NMP would aid development of novel cell specific therapeutics. This is an important step towards addressing an unmet need, namely therapeutic targeting of the inherent IRI in kidney transplantation, in an era of increasingly personalised medicine.

To address these questions, we used unbiased RNA transcriptomic analyses of human kidney biopsies taken at the end of NMP, to explore transcriptomic signatures of kidney IRI, correlated to clinical and histological outcomes. Furthermore, we used single-cell RNAseq data to deconvolute tissue transcriptomic signatures and outline the cellular landscape of severe IRI in injured kidneys undergoing NMP. We also isolated and characterised extracellular vesicles from the biofluids of *ex situ* perfused kidneys, revealing signatures reflective of processes active at the tissue level, for future exploration as potential biomarkers of graft injury.

## Methods

### Experimental design

To explore the transcriptomic signature of severe IRI during NMP, we used samples from a biobank of DCD kidneys perfused as part of the only randomised controlled trial of kidney NMP(9). We selected kidneys expected to have low IRI, given their immediate graft function after transplantation (primary function or haemodialysis for less than 24 hours post-transplant, n=10), and transcriptomically compared them to those hypothesised to have severe IRI, clinically manifested as more prolonged delayed graft function (haemodialysis beyond 24 hours, n=10). In the absence of an external clinical cohort of kidneys that underwent NMP prior to transplantation, we then sought to validate findings in an external research cohort of NMP kidneys (initially accepted but subsequently declined for transplantation). This included 7 paired kidneys previously randomised in a 1:1 fashion to undergo perfusion with either red blood cell (RBC) based perfusate, or an acellular perfusate; the full analysis of these kidneys is beyond the scope of this study, but important details are provided below. Using the degree of acute tubular injury as a histological surrogate of IRI/DGF(11–14), the kidneys were assessed and grouped into low (n=6) and high (n=7) acute tubular injury cohorts, which were histologically similar to IGF and PDGF kidneys respectively (discussed below), comprising the validation cohort.

### Clinical NMP kidney trial cohort

To further explore the tissue transcriptome of PDGF during NMP, samples from a biobank procured as part of the aforementioned RCT(9), were utilised (FigS1A). For bulk RNAseq, tissue biopsies taken at the end of NMP were selected for 10 IGF and 10 PDGF kidneys. Following total RNA extraction and quality assessment, nine samples (IGF n=6, PDGF n=3) yielded suitable RIN values (≥6) and were sequenced on an Illumina NovaSeq6000 platform. Following post-sequencing quality control, 7 samples (IGF n=4, PDGF n=3) were suitable for analysis. One further PDGF sample was excluded based on poor graft function explained by severe acute cellular rejection on day 7 post-transplant. Samples undergoing bulk RNAseq that had RNA remaining after sequencing were selected for small RNA seq; a total of 6 samples (IGF n=4, PDGF n=2) were sequenced at small RNA level. Following post-sequencing quality control (QC; discussed later) a total of 5 small RNAseq tissue samples were suitable for downstream analysis (IGF n=4, PDGF n=1). For extracellular vesicle (EV) small RNAseq analysis, paired urine and perfusate samples were identified for the same 10 IGF and 10 PDGF kidneys selected for tissue bulk RNAseq; RNA was isolated from a total of 20 urine (IGF n=10, PDGF n=10) and 20 perfusate (IGF n=10, PDGF n=10) derived EV samples. Following RNA quality assessment using Bioanalyser (discussed later), 33 samples had sufficient quality and quantity of RNA to proceed with sRNAseq. After post-sequencing QC, 7 perfusate (n=5 IGF, n=2 PDGF) and 15 urine (n=10 IGF, n=5 PDGF) EV RNA samples were suitable for downstream analysis. The final cohorts included for analysis are summarised in FigS1A: 6 samples for tissue bulk RNAseq (n=4 IGF, n=2 PDGF), 5 samples for tissue sRNAseq, 7 samples for perfusate EV sRNAseq (n=5 IGF, n=2 PDGF) and 14 samples for urine EV sRNAseq (n=10 IGF, n=4 PDGF).

#### NMP kidney trial protocol and definitions

Ethical approval for the previously completed trial(9) and collection of biological samples was granted by the East of England Cambridge Central Research Ethics Committee (15/EE/0356). Approval was specifically granted for a multicentre UK based open label randomised controlled trial of NMP in DCD kidney transplantation(9). Trial Registration Number: ISRCTN15821205. Recruitment was open to eligible patients receiving a controlled DCD kidney transplant (Maastricht Categories III & IV). All kidneys were procured following a National Organ Procurement Service protocol. Patients were randomised in a 1:1 fashion to receive a kidney either statically cold stored (SCS, n=200) or SCS followed by one hour of NMP (n=200). In the current study, only a subsample of the NMP arm was available from the remaining biobank. Delayed graft function was defined as the need for haemodialysis within the first 7 days post-transplant. However, previous work(10) showed no transcriptomic difference between kidneys experiencing immediate graft function (IGF) and those labelled as DGF due to haemodialysis within the first 24hrs (most commonly due to hyperkalaemia). Therefore, this study defined IGF as either primary function or haemodialysis for <24hrs post-transplant, and prolonged delayed graft function (PDGF) as dialysis required beyond 24 hours post-transplant.

#### NMP kidney trial perfusion protocol

The full perfusion protocol for RBC based NMP used in the RCT was published in Hosgood *et al*(9). In brief, after a period of SCS on ice at 4°C, the kidneys were weighed and prepared for perfusion. Renal artery and ureter were cannulated, and kidneys flushed with 1L of cold Ringer’s solution to remove preservation solution. Kidneys were then perfused using an adapted paediatric cardiac bypass system (Medtronic, Bioconsole 560). The circuit was primed with 300ml Ringer’s solution (Baxter Healthcare, Thetford, UK), 15ml Mannitol 10% (Baxter Healthcare), 27 ml sodium bicarbonate 8.4% (Fresenius Kabi, Runcorn, UK), 3000iu heparin (LEO Pharma A/S, Ballerup, Denmark) and 6.6mg Dexamethasone (Hameln Pharmaceuticals, Hamelin, Germany). A unit of ABO compatible packed red cells was then added. The perfusion solution was oxygenated (95% oxygen/5% CO_2_) at a flow rate of 0.1L/min and warmed to 35.5 - 36.5°C. Perfusion was undertaken continuously through the renal artery at a mean arterial pressure of 85mmHg and pump speed of 1450RPM. Nutrients (Synthamin 17 10%, Baxter Healthcare, Thetford, UK) and 15mls of sodium bicarbonate 8.4% (B Braun, Melsungen, Germany) were added. Insulin (100IU; Actrapid, Novo Nordisk, London, UK) was infused at a rate of 20ml/h, in addition to glucose 5% (Baxter Healthcare), at a rate of 5ml/h and Ringer’s solution was used to replace urine output (ml for ml).

### Research kidney NMP validation cohort

An external cohort of available machine perfused kidneys were used as a validation cohort. Seven pairs of kidneys procured for transplant but subsequently declined and offered for research were randomly assigned to perfusion with either a red blood cell (RBC) based perfusate, or an acellular based perfusate. All kidneys were procured following a National Organ Procurement Service protocol and transported under SCS. Ethical approval for the study was granted by the national ethics committee in the UK REC (22/WA/0167).

#### Machine perfusion within the validation cohort

Following SCS kidneys were weighed and prepared for perfusion in the same fashion as those from the kidney NMP trial above. The same perfusion system as above was used for both RBC and acellular perfused kidneys, however both were perfused at 32°C for six hours. RBC perfused kidneys received perfusate as per the NMP RCT. Acellular perfused kidneys were perfused with a Human serum albumin (5%, 250ml) and Ringer’s solution (219ml) based perfusate, supplemented with 6.6mg dexamethasone (Hameln Pharmaceuticals, Hamelin, Germany), 5ml calcium gluconate 10% and 15ml sodium bicarbonate 8.4% (Fresenius Kabi, Runcorn, UK), 250mg meropenem and 2.5mg verapamil. During perfusion Synthamin 17 10% (Baxter Healthcare, Thetford, UK) supplemented with 15mls of sodium bicarbonate 8.4% (B Braun, Melsungen, Germany), 5ml multivitamins, and 1ml insulin (100IU; Actrapid, Novo Nordisk, London, UK) was infused at 10ml/hr. Glucose 5% (Baxter Healthcare), at a rate of 3ml/h, and Glyceryl trinitrate (10ml of 50mg/ml in 90ml Saline) at 10ml/hr for the first hour then 5ml/hr thereafter, were also infused. Ringer’s solution was used to replace urine output (ml for ml).

#### Sample processing for bulk RNAseq analysis

In the validation cohort, RNA was extracted from tissue biopsies taken from seven kidney pairs at the end of SCS prior to commencement of NMP and at the end of 6 hours of NMP. All samples had a RIN value of ≥6 and proceeded to sequencing. Following QC, a total of 25 samples were suitable for downstream analysis (pre-NMP n=12; 6hrs NMP n=13, FigS1B).

### Histological assessment

Histological slides of all IGF (n=4) and PDGF (n=2) kidneys from the NMP trial cohort included in the bulk RNAseq analysis, were reviewed by a blinded specialist transplant pathologist (AP). We hypothesised that kidneys with IGF were more likely to have lower distribution and severity of acute tubular injury (ATI), and PDGF kidneys more likely to have wider distribution and severity of ATI; blinded review of histology confirmed this, with IGF kidneys better preserved in terms of ATI than PDGF kidneys. Blinded review of histology at the end of 6 hours of NMP in the validation cohort saw each kidney graded by distribution and severity of ATI, and the cohort ranked from lowest to highest ATI. The cohort was divided at the midpoint into low (n=6) and high (n=7) ATI cohorts. This resulted in a validation cohort with low ATI, histologically similar to IGF kidneys, and high ATI, histologically similar to PDGF kidneys.

### Sample storage prior to processing

Tissue biopsies were stored in RNAlater solution (Invitrogen RNAlater Solution) or snap frozen prior to transfer to -80°C until processing. For kidneys from the NMP RCT, perfusate and urine samples were also taken at the same time as tissue biopsies; samples were centrifuged at 1600rpm for 10min at 4°C, the supernatant (bar 200uL above the buffy coat) was collected and snap frozen in liquid nitrogen then stored at -80°C until analysed.

### Tissue RNA extraction

Samples were thawed on ice, removed from storage solution (if applicable), weighed, and then macerated on ice using a sterile scalpel. Macerated tissue was transferred to 700uL QIAzol Lysis Reagent (Qiagen, Hilden, Germany) and homogenised using a sterile pestle followed by further homogenisation via 10x passages through a 20g sterile needle. 140uL of chloroform was added and sample shaken for 15 seconds by hand before being centrifuged at 12,000g for 15 minutes at 4°C. The clear upper supernatant containing RNA was taken forward and 1.5x the volume of 100% ethanol was added. This was then loaded onto Qiagen RNeasy Midi columns and total RNA extracted according to manufacturer guidance. RNA was eluted in 50uL of RNAse free water. DNA was removed post-RNA extraction using TURBO DNA-free^TM^ kit (Invitrogen^TM^). Quality of RNA was assessed using nanodrop spectrophotometer and an RNA nano Bioanalyzer kit (Agilent Technologies, Palo Alto, CA, USA) using a Bioanalyzer 2100 (Agilent Technologies, Palo Alto, CA, USA). Samples with RIN >6 were sent for sequencing.

### Extracellular vesicle RNA extraction

Following SEC isolation of EVs from perfusate and urine as described below, RNA was extracted from EVs using the Plasma/Serum Exosome Purification and RNA Isolation kit (Norgen Biotek, Ontario, Canada) according to manufacturer instructions. Briefly, 600uL sample was mixed with recommended volumes of Lysis Buffers A and B before briefly vortexing, then incubating at room temperature for 10 minutes. Following incubation, 500uL of 96% ethanol was added to the mixture. Each sample was loaded on to the column 500uL at a time and centrifuged for 1 minute at 3000g. The column was then washed twice with Wash Solution A and then dried, spinning at 13,000g for one minute.

RNA was subsequently eluted in 40uL of RNAse free water and 1uL taken for quantification using a Bioanalyser Small RNA Analysis kit (Agilent Technologies, Palo Alto, CA, USA) on a 2100 Bioanalyser system (Agilent Technologies, Palo Alto, CA, USA).

### Bulk mRNA sequencing

RNA was shipped on dry ice to Genewiz Azenta (Germany) for RNA sequencing. Once received, RNA samples were quantified using Qubit 4.0 Fluorometer (Life Technologies, Carlsbad, CA, USA) and RNA integrity was checked with RNA Kit on Agilent 5300 Fragment Analyzer (Agilent Technologies, Palo Alto, CA, USA). rRNA depletion was performed using QIAGEN FastSelect rRNA HMR Kit (Qiagen, Hilden, Germany). RNA sequencing library preparation was performed with NEBNext Ultra II RNA Library Preparation Kit for Illumina following the manufacturer’s recommendations (NEB, Ipswich, MA, USA). Briefly, enriched RNAs were fragmented for 15 minutes at 94 °C. First strand and second strand cDNA were subsequently synthesized. cDNA fragments were end repaired, adenylated at 3’ends, and universal adapters were ligated to cDNA fragments, followed by index addition and library enrichment with limited cycle PCR. Sequencing libraries were validated using the Agilent Tapestation 4200 (Agilent Technologies, Palo Alto, CA, USA), and quantified using Qubit 2.0 Fluorometer (ThermoFisher Scientific, Waltham, MA, USA) as well as by quantitative PCR (KAPA Biosystems, Wilmington, MA, USA).

The sequencing libraries were multiplexed and loaded on the flowcell on the Illumina NovaSeq 6000 instrument according to manufacturer’s instructions. The samples were sequenced using a 2x150 Pair-End (PE) configuration v1.5. Image analysis and base calling were conducted by the NovaSeq Control Software v1.7 on the NovaSeq instrument. Raw sequence data (.bcl files) generated from Illumina NovaSeq was converted into fastq files and de-multiplexed using Illumina bcl2fastq program version 2.20. One mismatch was allowed for barcode matching.

### Small RNA sequencing

RNA library preparation and sequencing was conducted at Azenta Life Sciences (South Plainfield, NJ, USA). RNA samples were quantified using Qubit 2.0 Fluorometer (ThermoFisher Scientific, Waltham, MA, USA) and RNA integrity was checked with 4200 TapeStation (Agilent Technologies, Palo Alto, CA, USA). Small RNA sequencing libraries were prepared using NEB Small RNA library Prep Kit (New England Biolabs, Ipswich, MA, USA). In brief, Illumina 3’ and 5’ adapter was added to RNA molecules with a 5’-phosphate and a 3’-hydroxyl group sequentially. Reverse transcription was used to create single stranded cDNA. The cDNA was then PCR amplified using a common primer and a primer containing index sequence. Amplified cDNA construct was purified by polyacrylamide gel electrophoresis. The cDNA was concentrated by ethanol precipitation to deliver the final sequencing library. The sequencing library was validated on the Agilent TapeStation (Agilent Technologies, Palo Alto, CA, USA), and quantified by using Qubit 2.0 Fluorometer (ThermoFisher Scientific, Waltham, MA, USA) as well as by quantitative PCR (KAPA Biosystems, Wilmington, MA, USA). The sequencing libraries were multiplexed and clustered onto a flow cell on the Illumina NovaSeq instrument according to manufacturer’s instructions. The samples were sequenced using a 2x150bp Paired End (PE) configuration. Image analysis and base calling were conducted by the NovaSeq Control Software (NCS). Raw sequence data (.bcl files) generated from Illumina NovaSeq were converted into fastq files and de-multiplexed using Illumina bcl2fastq 2.20 software. One mismatch was allowed for barcode matching.

### Transcriptomic analysis

#### Bulk mRNAseq

The raw mRNAseq samples were subjected to preprocessing comprising subsampling without replacement(15) to 75M reads and trimming forward and reverse reads to 100 nucleotides (nts), (16) which reduced adapter contamination to within accepted levels (<5% across samples). The proportion of retained biological signal, before and after each processing step, was assessed using fastQC(17), summarised using multiQC(18). Poor quality samples, from a technical QC perspective, included two IGF samples with significantly higher overall GC% *(p*=0.025, under a standard t-test), calculated across the retained reads, as well as low numbers of unique reads indicating degradation; the mapping was performed using STAR (v2.7.4a)(19), with default parameters against vHg38 of the *H sapiens* genome. MultiQC was used to aggregate the quality reports from the fastQC, alignment and gene quantification, with ∼85% reads mapping uniquely across samples. The quantification of genes, based on the *H Sapiens* gtf annotation was performed using featureCounts(20). The two samples with poor RNA QC retained their poor assessment throughout mapping and quantification and were excluded from downstream analyses. An interactive bulkAnalyseR app(21) was subsequently generated on the retained samples; noise correction at expression-matrix level was performed using *noisyR* (22); the normalisation of gene expression was performed using quantile normalisation(23).

Differential expression analysis was performed using *DESeq2*(24)and *edgeR*(25). A log_2_(FC) threshold cut off of 0.75 and adjusted p-value 0.05, using Benjamini-Hochberg multiple testing correction, were used to determine DE mRNA transcripts for subsequent analyses. For gene set enrichment analysis (GSEA), all differentially expressed genes (log_2_(FC) threshold 0) were ranked by the inverse of the p value with the sign of the log fold-change, then ran against the Hallmark’s database within MSigDB(26), using the GSEA tool from the Broad Institute(27) with the pre-ranked option, and the following parameters: *No_collapse, classic, default settings*. Over-representation analysis was performed using *gProfiler2*(28), with ranked genes ran against the Gene Ontology, Kyoto Encyclopedia of Genes and Genomes (KEGG)(29) and Reactome Pathway (REAC)(30) databases, using all genes expressed in the deriving samples as background set. Single sample GSEA (*ssGSEA*) was carried out using normalized counts from quantile normalization using the GenePattern platform(31).

#### Bulk small RNAseq

Initial quality checking of the fastq files was performed using fastQC (17) and summarised using multiQC (18). The universal adapters were trimmed using custom scripts, on perfect matching against the first 8nts of the adapter sequence (GATCGTCG). Further quality checks(32) relayed on the distributions of redundant/non-redundant reads, complemented with complexity distributions (33). The mapping was performed using PatMaN(34), on 0 mis-matches and 0 gaps against version GRCh38 of the *H Sapiens* genome; the allocation of reads to annotation classes was performed against the *H Sapiens* gtf [classes explored in detail include miRNAs (35), piRNAs(36), tRNA halves (37)]. The expression level per stable fragment was defined as the number of identical redundant fragments; the expression level per annotation class was defined as the algebraic sum of redundant counts for reads perfectly incident with the annotation. For all sRNA analyses, performed at unique sequence level and annotation class, respectively, differential expression analysis was performed using *edgeR*(25) within *bulkAnalyseR*(21); a log_2_(FC) threshold cut off of 0.75 and adjusted p-value 0.05, using Benjamini-Hochberg multiple testing correction, were used to determine DE sRNAs for subsequent analyses. For miRNA analysis, functional annotation was performed using *TAM2.0*(38) with default settings, masking cancer related datasets. All analyses were performed in *R* (version 2023.12.1+402).

### Extracellular vesicle isolation and characterisation

#### Size exclusion chromatography

After thawing, perfusate and urine samples were centrifuged at 2500g for 15min at 4°C and the supernatant (bar 100uL) was taken forward. Size exclusion chromatography (SEC) was performed using qEV single 70nm SEC columns (Izon Science Ltd., Burnside, New Zealand). Prior to use, the 20% storage solution was allowed to run from the column and columns prewashed with 5mls of PBS. Sample was concentrated to 150uL using Amicon Ultra centrifugal ultrafiltration columns (100 kDa cut off, #UFC810024, Millipore, Burlington, MA, USA) and loaded directly onto the column, which was topped up with PBS after passage of the sample through the frit. The following previously validated fractions were collected: void fraction (Fraction 1, 1ml; subsequently discarded), intermediate/early EV fraction (Fraction 2, 600uL), expected EV rich fraction (Fraction 3, 600uL) and EV plus protein fractions (Fractions 4-6, 600uL)(39). Fractions were stored at -80°C before subsequent analysis.

#### Nanoparticle tracking analysis

Particle size and concentration were measured using a NanoSight NS300 (Malvern, Worcestershire, UK). A 500uL aliquot of sample was injected into the sample chamber using the Malvern NanoSight syringe pump system to maintain a constant flow rate. For all recordings, the camera level was set to 15, detection threshold to 5 and all other settings as default. Three 60s videos were recorded for each sample.

#### Western Blotting

Samples were mixed with 4x Laemmli Sample Buffer (#1610747, BIO-RAD, Hercules, CA, USA) and denatured for 5 minutes at 95°C. A total of 30uL sample was separated using a 4-20% SDS-PAGE (Mini-PROTEAN^®^ TGX Stain-Free™ Protein gels, #4568094, BIO-RAD, Hercules, CA, USA) and TBS running buffer. Transfer to nitrocellulose membranes was by semi-dry transfer using the BIO-RAD Trans-Blot SD transfer system. Membranes were blocked for one hour (TBS, 0.1% Tween, 3% dried milk). Primary antibodies against syntenin-1 (ab133267), ALIX (ab186728) and TSG101 (ab83), were incubated at 1:1000 dilution overnight at 4°C under continuous agitation. Corresponding horseradish peroxidase (HRP)-conjugated secondary antibodies (goat anti-rabbit (CST-7074), horse anti-mouse (CST-7076)) were incubated at 1:2000 concentration at room temperature for one hour under agitation. Membranes were then washed three times (TBS, 0.1% Tween) and chemiluminsence was activated using the Clarity Max Western ECL Substrate (#1705062, BIO-RAD, BIO-RAD, Hercules, CA, USA) and bands detected using the ChemiDoc Imaging System (BIO-RAD, BIO-RAD, Hercules, CA, USA).

#### Transmission Electron Microscopy

Transmission Electron Microscopy (TEM) was performed using a Tecnai G2 TEM Electron Microscope at the Cambridge Advanced Imaging Centre. Copper-carbon grids (400mesh, EM Resolutions, Sheffield, UK) were glow-discharged with argon using a Quorum K100X glow discharger. Grids were placed on a 5uL drop of pre-vortexed sample on dental wax for 1 minute. Grids were washed twice through transfer to drop of filtered distilled water and incubated for 30 seconds each. Excess fluid was then removed using filter paper. Staining was by grid transfer onto 5uL uranyl acetate (1.5% in distilled water) and incubated for 30 seconds before removal of excess fluid as before. Microscope was run at 200keV accelerating voltage using a 20um objective aperture. Images were acquired using an ORCA HR high resolution CCD camera via Hamamatsu DCAM board, running the Image Capture Engine, software version 600.323 (Advanced Microscopy Techniques, Danvers, USA).

### STRING analysis

Protein coding genes, identified based on their Ensembl IDs (ENSG) were converted to protein names and STRING(40) analysis undertaken. Two proteins were considered connected if they were part of the same physical subnetwork and had a mean interaction score of >0.7. Unconnected proteins were removed from the network and force direction graph plotted. The resultant network was subdivided using MCL clustering based on stochastic flow, and clusters annotated using selected GO terms associated with each cluster.

### Statistical analysis of clinical data

Mann-Whitney U and Wilcoxon rank sum tests were used to compare non-parametric continuous and ordinal demographic data, respectively. GraphPad Prism (version 9.0.2) was used.

## Results

The aim of this study was to investigate the transcriptomic signatures and cellular landscape underpinning IRI during human kidney NMP, using post-transplant kidney graft function/dysfunction as a clinical correlate of IRI severity. Delayed graft function, the clinical manifestation of IRI, is commonly defined as the need for haemodialysis within the first 7 days post-transplant(41). However, previous work(10) showed no transcriptomic difference between kidneys experiencing immediate graft function (IGF) and those with short term (<24hrs) DGF (haemodialysis commonly indicated for hyperkalaemia). Therefore, for transcriptomic analyses in this study, we defined immediate graft function (IGF) as primary function or requirement for dialysis within but not beyond the first 24 hours post-transplant. Prolonged DGF (PDGF), a clinical correlate of severe IRI, was defined as requirement for dialysis beyond the first 24 hours post-transplant.

The demographic characteristics of deceased-donors and the relevant clinical course post-transplant are presented in supplementary Table 1. Donors of kidneys that went on to develop PDGF were older (mean age: 67.5 (DGF) vs 45.5 (IGF), p=0.8847), predominantly male (100% vs 25%) and of similar BMI to IGF donors (median BMI: 31 vs 29, p=0.8444). Cold ischaemic time was longer in donors of PDGF kidneys (1132 mins vs 899 mins, p=0.3968), but warm ischaemic times were comparable to IGF kidney donors (45 mins vs 40 mins, p=0.7956). All grafts were functioning at one-year post-transplant, but 12-month eGFR was lower in the PDGF cohort (32.5ml/min vs 54ml/min, p=0.1349).

Quality control of bulk mRNAseq data is outlined in FigS2A-D. Initial transcriptomic comparison, applied on graft function, of IGF and PDGF kidneys across the width of the transcriptomic signature, revealed similar gene expression between IGF and PDGF kidneys (FigS2E); this was confirmed on principal component analysis (PCA; FigS2F); these findings are in line with expected transcriptomic characteristics given all kidneys underwent NMP following a period of SCS and are likely to share changes consistent with reperfusion. Heatmap representation of differentially expressed genes (DEGs) highlighted one PDGF kidney as transcriptionally dissimilar to the other PDGF kidneys, particularly in terms of upregulated genes, which better aligned to the transcriptional profile of IGF kidneys (FigS2G). Investigation of clinical data in the recipient of this kidney revealed the aetiology of PDGF to be acute T cell mediated rejection, confirmed with biopsy on day 7 post-transplant, with significantly improved graft function following treatment with methylprednisolone and immunosuppression modulation. Given graft dysfunction was likely due to a memory immune response, rather than a true DGF secondary to acute tubular injury (ATI), this kidney was excluded from the PDGF cohort to ensure those included in the analysis had PDGF underpinned by ATI and IRI. The final cohorts showed clear differences between IGF (n=4) and PDGF (n=2) on PCA (FigS2I), with each group sharing similar DEGs (Fig1A).

**Figure 1.**
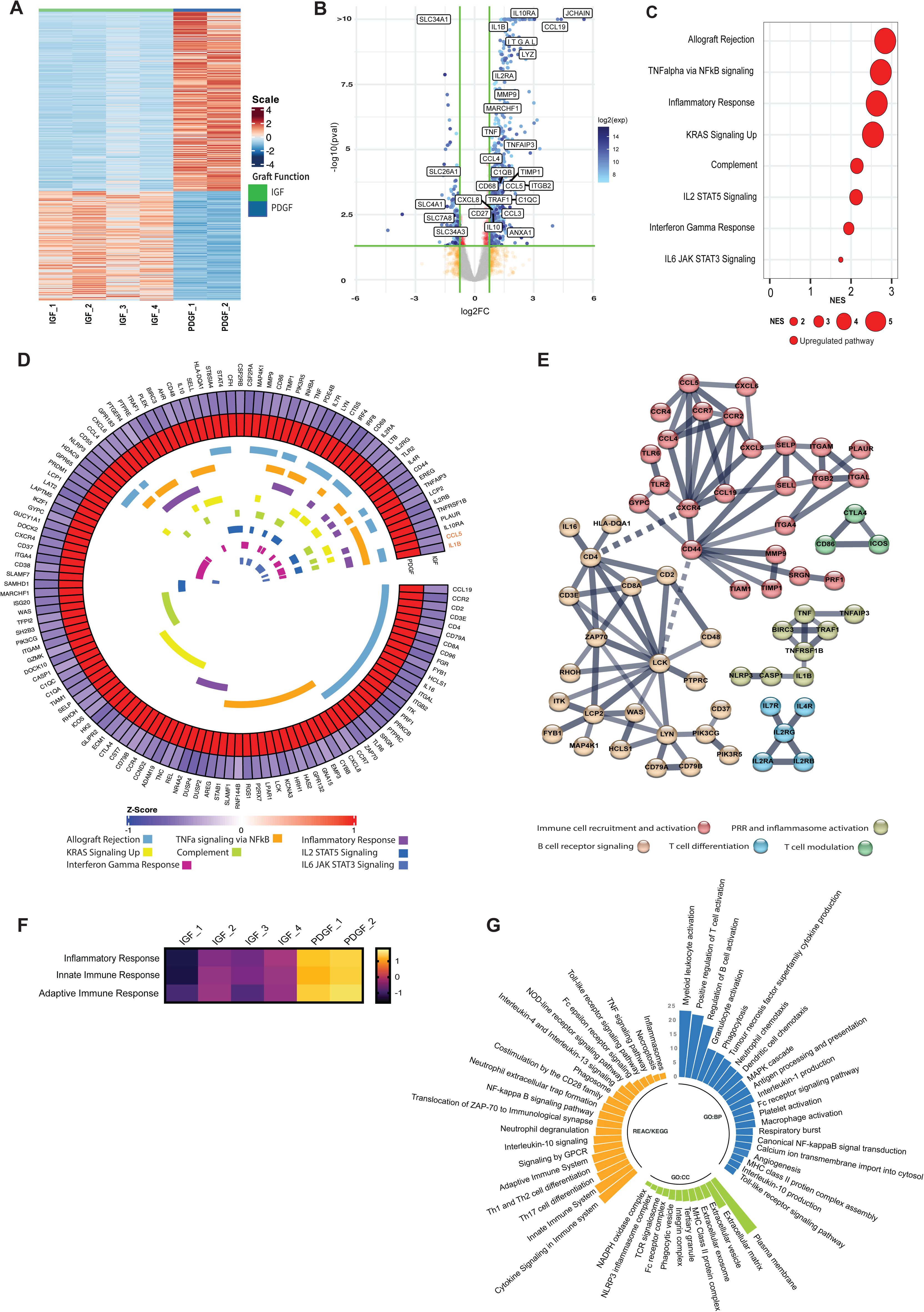
Bulk mRNA transcriptional analysis reveals immune and inflammatory pathway activation drives the core signature of prolonged delayed graft function. **A**: Heatmap demonstrating differentially expressed genes between IGF and PDGF kidneys (Supplementary Table 2). **B**: Volcano plot of genes differentially expressed in PDGF kidneys, annotated with upregulated pro-inflammatory and immune related genes, and downregulated solute transporter genes. Exp; expression. **C**: Gene Set Enrichment Analysis (GSEA) on differentially expressed genes (logFC1.5,p value<0.05) against the MSigDB Hallmarks dataset(26), in PDGF kidneys reveals immune and inflammatory pathway activation driving severe IRI (Supplementary Table 3). Only significant pathways are plotted (FDR <0.05). Size of bubble corresponds to degree of enrichment. NES, normalized enrichment score; FDR, false discovery rate. **D**: Circular heatmap of leading edge genes from enriched pathways in C; internal colours represent pathways each specific gene is annotated to; genes highlighted in orange (*CCL5, IL1B*) show greatest cross-pathway annotation (five pathways; Supplementary Table 4). **E**: STRING analysis of leading edge genes from C; solid line represents interaction within protein cluster, dashed line represents interaction between clusters. **F**: Heatmap of scaled enrichment of pathways from single sample GSEA. **G**: Bar plot of pathways over-represented in PDGF kidneys using significantly differentially expressed genes (Supplementary Table 5).

### Immune and inflammatory pathway activation drives the core signature of prolonged delayed graft function

Gene expression comparison identified 841 DEGs in PDGF compared to IGF kidneys, including 674 upregulated and 167 downregulated genes (Fig1B; Supplementary Table 2). Upregulated genes comprised those encoding pro-inflammatory cytokines (*TNF, IL1B, CXCL8/IL8),* immune cell recruiting chemokines (*CCL3, CCL4, CCL5*), integrin-mediated leucocyte adhesion molecules (*ITGB2, ITGAL*), and extracellular matrix remodeling proteins (*MMP9, TIMP1*). Genes associated with cells of monocytic lineage (*CD68, LYZ*, *CCL19*), T cells (*IL2RA, MARCHF1)* and B cells (*JCHAIN, CCL19*) were also upregulated. Upregulation of complement associated genes (*C1QB, C1QC*) was also seen in PDGF kidneys. Interestingly, genes associated with anti-inflammatory and immune regulation responses (*IL10, IL10R, TNFAIP3, CD27*) were also upregulated compared to IGF kidneys. Downregulated genes included those associated with tubular cell function, specifically in solute transport (*SLC4A1, SLC7A8, SLC26A1, SLC34A1, SLC34A3*).

Gene set enrichment analysis (GSEA) using all genes differentially expressed in PDGF kidneys recapitulated the enrichment of pro-inflammatory pathways previously suggested using kidneys from the same RCT(10), including *TNFa via NFkB signaling* and *Inflammatory response*, whilst simultaneously confirming depletion of key metabolic pathways, namely *Oxidative phosphorylation* (FigureS3). These findings validate the global transcriptomic signature previously shown in PDGF kidneys during NMP(10). Next, we performed GSEA using only protein coding genes that were significantly differentially expressed in PDGF kidneys, to establish a core transcriptional signature of severe IRI (Fig1C). This demonstrated persistent enrichment of eight immune and inflammatory pathways, including *Allograft rejectio*n, *TNF alpha signaling via NFkB* and *Inflammatory response* (Fig1C). Leading edge analyses of these eight pathways driving severe IRI identified 133 unique driver genes (Supplementary Table 4). When looking at overlapping gene expression between pathways (Fig1D), only two genes were part of 5 or more pathways: *CCL5*, an immune cell recruitment chemokine, specifically for CCR5-, CXCR3- and CXCR5-expressing lymphocyte subsets, and *IL1B*, a potent pro-inflammatory cytokine secreted by pro-inflammatory (M1) macrophages following inflammasome activation. STRING analysis of the leading edge genes (Fig1E) showed five major nodes: immune cell recruitment and activation (*CCL4, CCL5, CXCL8, CCR7, CD44, CD2, CD3E),* pattern recognition receptor (PRR) and inflammasome activation (*TNF, IL1B, NLRP3, CASP1, TNFAIP3*), B cell receptor signalling (*LYN, CD79a, DOCK2)*, T cell differentiation (*IL2RA, IL4RA, IL7RA)*, and T cell modulation (*CTLA4, CD86, ICOS*). Taken together, these results highlight a pro-inflammatory immune mediated injury underpinning severe IRI in kidneys developing PDGF, with both innate and adaptive immune response components. This signature is present after just one hour of NMP and may reflect a significantly up- or dys-regulated innate immune process, supported by leading edge genes that translate to cytokines/chemokines secreted by cells of myeloid lineage, and activation of TNFalpha and NFkB signaling pathways, both of which are essential components of innate immunity.

### Kidneys with prolonged delayed graft function show enrichment for innate immune processes driven by Nuclear Factor-kappa B signaling during NMP

Having established a core transcriptomic signature of severe IRI in PDGF kidneys, we sought to better characterise the underpinning mechanisms, employing over-representation analysis (ORA) querying the Gene Ontology, KEGG, and Reactome protein databases, to investigate the functional role of genes and related pathways driving the transcriptomic signature (Supplementary Table 5). PDGF kidneys were enriched for inflammatory response and both innate and adaptive immune responses, with many of the enriched pathways linked to these higher order terms in the gene ontology database (Fig1F). Kidneys with PDGF displayed enrichment of Gene Ontology terms associated with cytokine production and leucocyte chemotaxis, corroborating the STRING analysis. These kidneys were also enriched specifically for recruitment and migration of myeloid leucocytes, macrophage activation and phagocytosis, further evidence of innate immune activation in PDGF (Fig1G). Furthermore, we found enrichment of pro-inflammatory signaling, specifically through *MAPK cascade*, *Canonical NF-kappaB signal transduction*, and *Toll-like receptor signaling pathway*. Enriched cellular components suggested cell interaction through integrins, likely at the plasma membrane and with remodeling of the extracellular matrix (Fig1G). Innate immune function related cellular components such as *phagocytic vesicle*, *NLRP3 inflammasome complex* and *NADPH oxidase complex* were also enriched. Genes upregulated in PDGF were also annotated to extracellular vesicles (EVs), promising novel biomarkers and potential therapeutic/drug delivery systems in transplantation(42). Querying the KEGG and Reactome pathway databases further highlighted the role of NFkB signaling in PDGF (Fig1G), in addition to enrichment of PRR activation (*TLR signaling*, *NOD-like receptor signaling pathway*), a common signaling event initiating NFkB pathway activation. There was also enrichment in the downstream consequences of NFkB activation, including cytokine signaling, *Th17 cell differentiation* and *inflammasome activation*. Importantly, whilst we show the majority of pathways enriched in PDGF are pro-inflammatory, we identified enrichment of *Interleukin-10 signaling* and *Interleukin-4 and interleukin-13 signaling*, both of which are involved in resolution of the immune/inflammatory response, promoting anti-inflammatory/pro-repair ‘M2’ macrophage polarisation(43). Taken together, our results reveal a strong innate immune signature present during NMP in kidneys going on to suffer PDGF, including innate-adaptive immune crosstalk and propagation of inflammation.

### Mononuclear phagocytes are enriched in kidneys during NMP that develop prolonged delayed graft function

Having established NFkB signaling as a key upregulated pathway in kidneys with PDGF and an associated enrichment of innate immune system pathways, in addition to evidence of innate-adaptive immune crosstalk, we sought to infer the cellular landscape underpinning severe IRI during NMP in PDGF kidneys. We used curated cell-type signatures from a spatiotemporally resolved kidney atlas(44) (Supplementary Table 6) to deconvolute bulk RNAseq. The signature of four subtypes of mononuclear phagocytes (MNP), were found to be enriched in PDGF kidneys (Fig2A), including classical (MNPa) and non-classical (MNPb) monocyte derived phagocytes, typically of pro-inflammatory M1 macrophage phenotype(44). Enrichment of dendritic cells (MNPc) was also seen, in line with the enrichment of the *antigen processing and presentation* pathway identified in the ORA. Tissue macrophages (MNPd) were also enriched. Whilst macrophages are classically defined as M1 or M2, there is a spectrum of polarisation depending on the stimulus(45). To explore this in PDGF kidneys, we undertook GSEA using curated cytokine-specific macrophage polarisation state signature reference datasets(45) (Supplementary Table 8). We found pro-inflammatory M1 phenotype macrophage signatures to be enriched in this cohort, specifically those polarised by TNF, IFNgamma and Prostaglandin E2 (Fig2B). There was enrichment of IL4 polarised macrophages, typically displaying an anti-inflammatory M2 phenotype, however, to a much lesser extent than M1. In addition to MNPs, adaptive immune subsets were enriched, including CD8 and CD4 T cells, B cells, NK cells, highlighting the importance of adaptive immune cells in PDGF and innate-adaptive cross talk highlighted in the earlier ORA.

**Figure 2.**
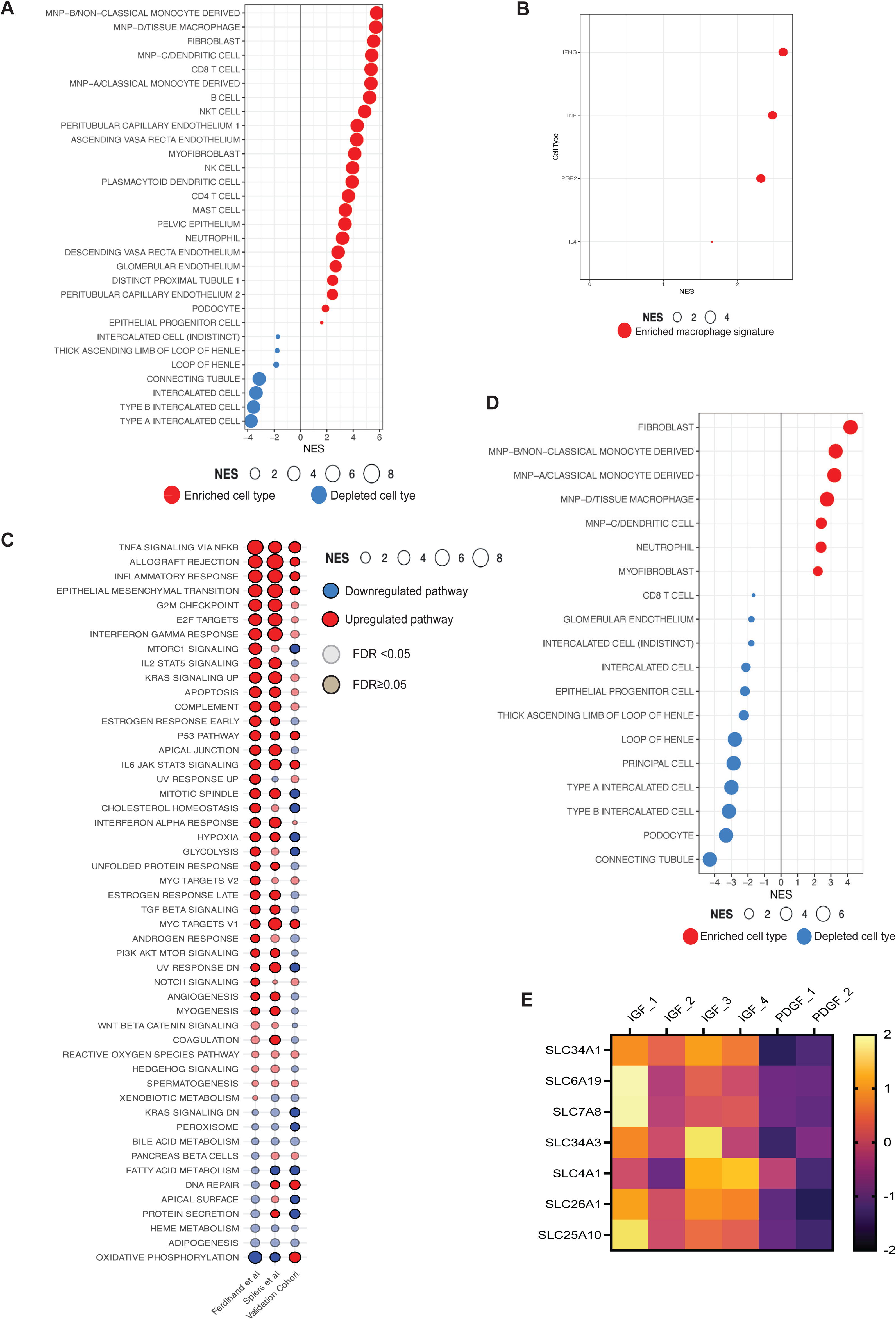
Cell expression signatures enriched in kidneys developing prolonged delayed graft function. **A**: GSEA of differentially expressed genes in PDGF using curated scRNAseq signatures of kidney cell types from a spatio-temporally resolved human kidney atlas (Supplementary Table 6,8)(44). Only significant cell types are plotted (FDR <0.05). Size of bubble corresponds to degree of enrichment. NES, normalized enrichment score; FDR, false discovery rate. **B**: GSEA of the same genes as A using cytokine-specific macrophage polarisation signatures (Supplementary Table 7). **C**: Comparison of GSEA against the Hallmarks datasets between kidney NMP datasets, including Ferdinand et al(10), contemporary analysis and validation cohort (Supplementary Table 3,8). Solid colour represents FDR <0.05, transparency represents FDR β0.05. **D**: GSEA of differentially expressed genes in high acute tubular injury kidneys from the validation cohort against curated scRNAseq signatures(44). Only significant cell types are plotted (FDR <0.05). Size of bubble corresponds to degree of enrichment. NES, normalized enrichment score; FDR, false discovery rate. **E**: Heatmap depicting expression of solute transporter genes between graft function types.

### Enrichment of innate immune cell signatures persists at six hours of machine perfusion

The previous analyses highlight the central role of the innate immune system, in particular monocyte/macrophage activation, in the pathogenesis of severe IRI and its clinical manifestation as PDGF. To validate this cellular landscape, an external cohort of machine perfused kidneys (initially procured for transplant but subsequently declined and offered for research) were identified (Supplementary Table 9). These kidneys were part of a study assessing kidney NMP using RBC compared to acellular perfusate; full analysis is beyond the scope of this study. However, at bulk transcriptomic analysis, PCA revealed similar gene expression patterns between RBC and acellular perfused kidneys, both in terms of change between pre-NMP and six hours (end) NMP timepoints, as expected when an organ goes from a state of relative metabolic inactivity to reactivation during perfusion, and without separation of samples at the six-hour timepoint (FigS4B). PCA highlighted some separability based on the gender of kidney donors (FigS4B), with donor sex subsequently included as a co-variate for differential expression. After six hours of NMP, only four genes were differentially expressed between RBC and acellular perfused kidneys, namely the haemoglobin related genes (HBA1, HBA2, HBB; FigS4C). Given the lack of significant transcriptomic differences between RBC and acellular perfused kidneys at 6 hrs of NMP, they were considered together as the validation cohort for this study.

Biopsies at the end of NMP in the validation cohort were assessed by a blinded specialist transplant histopathologist (AP) for degree of acute tubular injury (see Histology Assessment in supplementary methods)(46). Based on this histological grading, the validation cohort was divided into low ATI (n=6) and high ATI (n=7) groups. Low ATI kidneys were found to share histological similarities with IGF kidneys, and high ATI kidneys were histologically similar to PDGF kidneys in the clinical NMP trial cohort (FigS4A). Thus, the degree of ATI was used as a histological surrogate endpoint for IRI severity/PDGF. We performed bulk transcriptomic analysis of tissue biopsies taken at the end of 6 hours machine perfusion in the validation cohort, and compared gene expression signatures between low and high ATI kidneys.

High ATI compared to low ATI kidneys showed enrichment of *‘TNFalpha signaling via NFkB’, ‘Inflammatory response’* and *‘Allograft rejection’* pathways (Fig2C), recapitulating the clinically correlated IRI signature seen in PDGF kidneys in the NMP trial derivation cohort, and in a prior study from the same trial(10). Deconvolution of DEGs at 6 hours of NMP in high ATI kidneys (i.e. those sharing histological similarity with PDGF kidneys) revealed consistent enrichment for mononuclear phagocyte signatures (Fig2D), similar to those observed during NMP in kidneys with PDGF in the transplanted cohort. These findings provide further support for the role of inflammatory innate immune cells of myeloid lineage in the pathogenesis of IRI in severely injured kidneys during NMP, and indirectly validate the transcriptomic signature observed in NMP kidneys that went on to develop PDGF. The analysis also suggests that the innate immune cell signature present at one hour of NMP is sustained at six hours of NMP in severely injured kidneys.

### Kidneys with severe IRI/PDGF show increased expression of gene signatures associated with fibroblasts/myofibroblasts and depletion of renal tubular cell signatures during NMP

Kidneys suffering more severe IRI (PDGF kidneys and high ATI kidneys in the respective cohorts) also showed enrichment for non-immune cells, including fibroblasts and myofibroblasts (Fig2A,D). The global signature of PDGF (in both previous(10) and current PDGF cohorts) and in the high ATI cohort, included enrichment for *epithelial-mesenchymal transition* (EMT, Fig2C), shared by fibroblasts and myofibroblasts in kidney fibrosis(47).

We observed depletion of several renal tubular epithelial cell (TECs) signatures during NMP in kidneys that went on to develop PDGF (Fig2A). This was recapitulated in the validation cohort (Fig2D). Interestingly, genes downregulated during NMP in PDGF kidneys included those encoding solute carrier-mediated transmembrane transport proteins (Fig2E). Genes such as *SLC4A1* and *SLC7A8* are predominantly expressed in the tubuloepithelial cells, and *SLC26A1, SLC34A1 and SLC34A3* are specifically expressed in proximal convoluted tubular cells (48); both cell populations were depleted in PDGF kidneys. These genes are annotated to the ‘*transport of inorganic cations/anions and amino acids/oligopeptides*’ pathway (Reactome Database: R-HSA-425393), encompassing acid-base balance, glucose, and protein absorption in the tubuloepithelial compartment of the kidney. In agreement, ORA of significantly downregulated genes revealed depletion of this pathway in PDGF kidneys.

### RNA cargo of kidney-specific extracellular vesicles released during NMP

#### Small RNA cargo in extracellular vesicle perfusate and urine compartments

Our tissue bulk RNAseq analysis highlighted ‘*extracellular vesicle*’ and ‘*extracellular exosome*’ cellular components to be enriched in kidneys with severe IRI. Given EVs, nanosized lipid bilayer delimited particles known to contain a bioactive cargo of proteins, lipids and miRNAs(49), have shown promise as biomarkers and therapeutic targets in transplantation(42), we hypothesised that EVs released by kidneys into perfusate and urine compartments during NMP may harbour signatures of graft injury. We isolated EVs by size exclusion chromatography (SEC) with a previously validated method(39), and characterised them in accordance with international guidelines(50). Nanoparticle tracking analysis revealed particles in the SEC fraction expected to be EV rich (fraction 3) to have a mean and mode size in the small EV (sEV) range, with an average concentration of 1x10^9^ (FigS5A-B). Western Blot confirmed the presence of EV specific protein markers (syntenin-1 and ALIX; FigS5C) and transmission electron microscopy demonstrated typical sEV morphology in this fraction (FigS5D). These data confirm the presence of EVs in the urine and perfusate of kidneys undergoing NMP.

Next, we capitalised on the opportunity to study the small RNA cargo of kidney-specific EVs in our isolated *ex situ* organ perfusion system which avoids the perturbations of the *in vivo* human setting where EVs in biofluids (e.g. urine) originate from cells along the urogenital track. Following QC (FigS6), quantification of isolated EV RNA demonstrated significant variability in the proportion of perfusate EV (pEV) RNA in the miRNA size range (FigS5E) and a significantly lower ratio of miRNA/total sRNA compared to urine EVs (uEV). As expected, RNA species >200nt was highest in tissue samples where polyadenylated long RNA species predominate (FigS5F), with no difference between IGF and PDGF samples.

We examined miRNAs in tissue, uEV and pEV of IGF kidneys undergoing NMP, as they represent a healthier cohort of kidneys maintained in a near physiological state with warm oxygenated perfusate and nutritional supplementation. A total of 576 unique miRNAs were present across tissue (n=4), uEV (n=10) and pEV (n=5) compartments. PCA calculated on the top most abundant 200 miRNAs (by expression) highlighted linear separability between tissue and EV compartments (FigS5G). Some partial separation was also seen between urinary and perfusate EV miRNA profiles, with overlap likely attributable to a single outlying pEV sample. Our data on kidney-specific EV composition and RNA cargo have been deposited for potential use as a reference dataset in future studies (link in Data Statement).

#### Extracellular vesicles released during NMP may hold miRNA signatures of graft injury

Having demonstrated that EVs in different compartments contain a proportion of distinct miRNAs, we hypothesised they may also provide unique signatures related to the degree of graft injury and post-transplant function. First, we examined miRNAs in both uEV and pEV. In both groups, the top 30 most abundant miRNAs accounted for over 85% of all miRNAs (Fig3A&B). The most abundant miRNA in both EV compartments was hsa-let-7b-5p, a common Ago2-complex miRNA found in the kidney that may have a renoprotective role in attenuating renal fibrosis(51). This was followed by hsa-miR-148a-3p, a miRNA known to promote differentiation and M1 polarisation of macrophages(52), *in vitro*. The abundance of this miRNA in uEVs was similar between IGF and PDGF derived samples (Fig4A).

**Figure 3.**
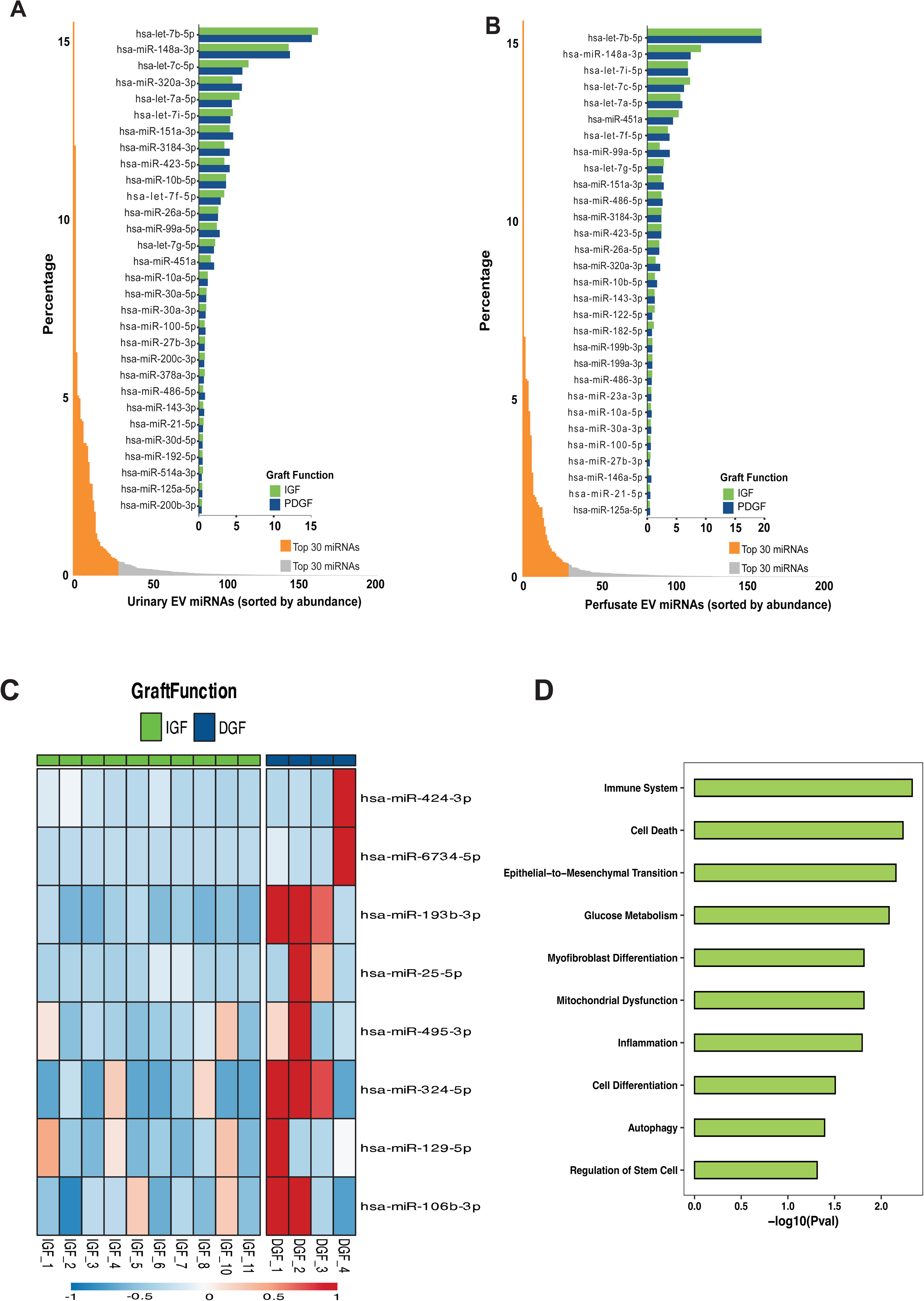
Extracellular vesicles in the biofluids of *ex situ* perfused kidneys harbor miRNA signatures reflecting tissue processes in the parent organ. Bar plots of urinary (**A**) and perfusate (**B**) EV miRNAs by abundance. Lower plots represent all miRNAs present in each source of EVs, with orange denoting the top 30 miRNAs by abundance. Upper plot illustrates top 30 miRNAs in urinary (**A**) and perfusate (**B**) EVs by graft function. **C**: Heatmap of the eight urinary EV miRNAs differentially expressed between IGF and PDGF kidneys. **D**: Functional annotation of the differentially expressed miRNAs in PDGF urinary EVs.

Next, we focused on uEVs given their ability to be non-invasively sampled in donor, recipient and during NMP, making them attractive biomarker candidates. Differential expression between IGF uEVs (n=10) and PDGF uEVs (n=4) identified eight upregulated miRNAs (Table 1). A heatmap summary showed no single miRNA to be uniformly upregulated in all PDGF samples (Fig3C). However, two miRNAs were upregulated in three of the four samples: hsa-miR-193b-3p and hsa-miR-324-5p. Both are associated with renal fibrosis, and hsa-miR-193b-3p directly associated with tubulointerstitial inflammation, a hallmark of kidney IRI(53). Functional annotation of the differentially expressed uEV miRNAs in PDGF demonstrated enrichment in processes associated with the immune system, EMT and inflammation, in addition to myofibroblast differentiation (Fig3D). Taken together, these data highlight the potential of uEVs to harbor miRNA signatures reflective of renal pathology, requiring further examination as potential biomarkers of IRI during NMP.

**Table 1.** Differentially expressed miRNAs in urinary EVs from prolonged delayed versus immediate graft function kidneys.

| miRNA ID | Function | log2FC | Adjusted P value |
| --- | --- | --- | --- |
| hsa-miR-6734-5p | Inhibits the growth of colon cancer cells by up-regulating p21 gene expression and subsequent induction of cell cycle arrest and apoptosis(67). | 5.80 | 1.40E-07 |
| hsa-miR-424-3p | Promotes stem-like features and tumorigenic capability in prostate cancer via EV transfer(68). | 3.99 | 0.00185004 |
| hsa-miR-193b-3p | Associated with tubulointerstitial inflammation and fibrosis and predicts development of chronic kidney disease after radical nephrectomy in patients without prior nephropathy(53). | 3.25 | 1.27E-08 |
| hsa-miR-129-5p | Protects against neuronal IRI by inhibiting HMGB1 and TLR3-associated cytokines *in vitro*(69). | 2.49 | 0.0490998 |
| hsa-miR-495-3p | Potential regulator of cellular reactive oxygen species during hypoxia(70). | 2.42 | 0.01486829 |
| hsa-miR-106b-3p | Expressed in macrophages from human kidneys suffering DGF(71). Upregulated in uEVs of paediatric kidney transplant recipients with subclinical rejection(72). | 2.12 | 0.0490998 |
| hsa-miR-324-5p | Promotes renal fibrosis in murine models(73,74). | 1.65 | 0.00378879 |
| hsa-miR-25-5p | Protective against diabetic nephropathy in murine models(75). Reduces pro-inflammatory cytokine levels following treatment with lipopolysaccharide in murine models(76). | 1.40 | 0.0490998 |

## Discussion

Ischaemia-reperfusion injury, which remains an unmet need in renal transplantation, manifests as graft dysfunction following implantation that, in severely injured grafts, can be prolonged and lead to increased graft immunogenicity and to poorer long-term outcomes. We dissected the mechanisms of IRI during *ex situ* organ NMP, characterising the transcriptomic differences between IGF and PDGF kidneys, capitalising on a unique biobank derived from the first, and currently only, clinical RCT in kidney normothermic machine perfusion. Having validated global pro-inflammatory signatures associated with development of PDGF following kidney NMP(10), we went further and identified a core transcriptional signature of severe IRI. This included transcriptional pathways corresponding to *‘TNFalpha signaling via NFkB’, ‘Allograft rejection’* and *‘Inflammatory response’*. These molecular processes were underpinned by activation of the innate immune system, innate-adaptive immune crosstalk and propagation of inflammation in severe IRI. Indeed, deconvolution of bulk RNA sequencing using scRNAseq signatures showed enrichment of proinflammatory mononuclear phagocytes, fibroblasts and myofibrolasts and relative depletion of tubular epithelial cell signatures in kidneys with severe IRI and prolonged DGF. These findings were recapitulated in perfused kidneys with high versus low acute tubular injury, correlated histologically with PDGF and IGF kidneys respectively, supporting further the core transcriptional signature and cellular landscape underpinning severe IRI. Finally, we performed, to our knowledge, the largest study characterising kidney-specific EVs released in the perfusate and urine compartments in the setting of isolated *ex situ* kidney NMP and provide preliminary evidence that urinary EVs from kidneys with PDGF are enriched with miRNAs associated with inflammatory and pro-fibrotic processes, potentially reflecting the tissue microenvironment of kidneys with severe IRI.

NMP provides a unique platform for multiparametric assessment of kidneys pre-transplant(8). Integrating transcriptomics data could enable early assessment and identification of grafts that could benefit from intervention. *Ex situ* organ NMP also provides an excellent platform for the delivery and testing of therapeutics. Our validated transcriptomic signature of severe IRI in kidneys during NMP could provide a useful endpoint for assessment of therapeutic efficacy *ex situ*. We found significant enrichment of genes and pathways associated with activation of the innate immune system during NMP in both PDGF kidneys and in kidneys with severe ATI. The innate immune response plays a key role in kidney injury(54), with signaling via NFkB mediating many of the relevant cellular responses and the propagation of inflammation(55). In kidney IRI recapitulated during NMP, we saw strong upregulation of NFkB signaling and associated pro-inflammatory downstream mediators (e.g. IL1B, TNFa, CXCL8/IL8), as observed in other organ systems(56,57), highlighting NFkB signaling as a therapeutic target in solid organ transplantation. Many approaches to NFkB inhibition have been trialed(58), but systemic administration is compromised by off-target effects on various physiological responses, and multiple cell types, making translation challenging. Targeting NFkB during NMP removes the risk of systemic toxicity and off-target effects(59,60), and could be enhanced by targeting of key cells implicated in IRI (61).

Using scRNAseq to deconvolute bulk RNAseq, we demonstrated cell-specific expression signatures during kidney NMP for the first time. PDGF kidneys had significant enrichment for pro-inflammatory innate immune cell signatures, including MNPs consistent with M1 macrophages, a finding also present in the independent cohort of NMP kidneys with high ATI, which were histologically similar to PDGF kidneys. Our findings provide evidence for the role of inflammatory innate immune cells of myeloid lineage in the pathogenesis of IRI in severely injured kidneys. Interestingly, the innate immune cell signature present at one hour of NMP was also present after six hours of NMP in severely injured kidneys. In kidneys with poor long-term function (low eGFR), which correlates with severe IRI and prolonged DGF, biopsies taken immediately post-retrieval demonstrated enrichment of tissue macrophage, NK cell and adaptive immune cell signatures(57), observed in both cohorts of more severe IRI in our study. This may suggest activation of the innate and adaptive immune cell populations starts at the time of donor organ procurement, particularly in marginal donor kidneys, and is further exacerbated during reperfusion, leading to severe IRI. Importantly, tissue kidney biopsies at the time of organ procurement in allografts with low 12-month eGFR (57), showed enrichment for fibroblasts/myofibroblasts and EMT, similar to the enrichment we observed in kidneys during NMP that develop PDGF or with severe ATI (validation cohort). This observation suggests the process of pro-fibrotic gene induction could start at the time of organ procurement and, in the context of the pro-inflammatory pathway upregulation and dysregulated innate immune activation seen upon reperfusion, impaired healing may be sustained in kidneys suffering severe IRI, leading to poor long-term outcomes.

We acknowledge an important limitation of this unbiased exploratory study is the small number of kidneys in the PDGF cohort. This reflects the challenges with biobanking in a real-world clinical setting and the need to support multiple translational studies. To address this limitation, in the absence of additional samples to increase the cohort, we relied on the convergence and robustness of bioinformatics analyses(25) and on internal variation assessment approaches such as the weighted likelihood empirical Bayes method. This statistical approach achieves highly stable and replicable results even for experiments with very small numbers of biological replicates(64), as reflected in the fact we were able to confirm previously detected global transcriptomic signatures associated with PDGF that were derived from a larger subcohort of the same RCT(10). We were then able to replicate the transcriptomic and cell expression signatures established using this method in an external validation cohort, of non-transplanted NMP kidneys with high ATI (histologically similar to PDGF kidneys). The validation cohort contained a larger number of samples, highlighting the reproducibility of findings and providing confidence in the results obtained in the smaller derivation cohort.

A novel observation in the current study was the increased expression of genes related to EV cellular components in kidneys during NMP. We, therefore, isolated EVs from the biofluids of kidneys during NMP and provided the largest, international guideline compliant(50), characterization of EVs released in an isolated organ perfusion system. Subsequent unbiased transcriptomic analysis revealed EV miRNA biosignatures reflective of tissue processes seen in severely injured kidneys during NMP, that require further investigation. MicroRNA signatures have been extensively studied and pursued as biomarkers of renal disease (65,66). However, these do not necessarily reflect renal specific pathology, given many patient-derived biofluids (e.g. peripheral blood and urine) contain EVs released from other organs. The isolated organ system provided by NMP is a ‘clean’ system, allowing organ-specific EVs to be investigated. EVs isolated from this system hold excellent biomarker potential, given their bioactive cargo is representative of the physiological state of the parent tissue. Accordingly, our deposited data on kidney-specific EV composition and RNA cargo provides a much-needed resource (e.g. miRNA reference dataset) for future studies.

In summary, we provide a core transcriptomic signature of IRI during NMP in kidneys going on to develop PDGF, enriched for innate immune processes. We have identified the cell expression landscape of kidneys suffering severe IRI during NMP, including cell expression signatures enriched for pro-inflammatory immune cells, fibroblasts and myofibroblasts and depleted for tubuloepithelial cells. We have also highlighted the potential for kidney-specific EVs to serve as biomarkers of tissue pathology during NMP, worthy of exploration in future studies.

## Data Availability

All transcriptomic data produced within this study will be made available via GEO Accession Number upon publication of the manuscript.

## Acknowledgements

We acknowledge funding support from National Institute for Health and Care Research (NIHR) Blood and Transplant Research Unit in Organ Donation and Transplantation (NIHR203332), a partnership between NHS Blood and Transplant, University of Cambridge and Newcastle University. The views expressed are those of the authors and not necessarily those of the NIHR, NHS Blood and Transplant or the Department of Health and Social Care. HS acknowledges funding from a Sir Roy & Lady Calne Royal College of Surgeons Research Fellowship (G121988). IM acknowledges funding by the Wellcome Trust [203151/Z/16/Z] and the UKRI Medical Research Council [MC_PC_17230]. VK acknowledges funding from an NIHR Fellowship (PDF-2016-09-065) and as a Paul I. Terasaki Scholar (G106170). We thank Professor Tim Williams (Department of Veterinary Medicine, University of Cambridge) for use of the Nanoparticle Tracking Analyser. This research was supported by the Cambridge Advanced Imaging Centre with thanks to Dr Karin Müller and Dr Filomena Gallo. For the purpose of open access, the authors applied a CC BY public copyright license to all versions of the manuscript arising from this submission.

## Data Statement

The sequencing data and metadata obtained from samples of the randomised controlled trial(10) that support the findings of this study will be made available upon publication. Data from the validation cohort will be made publicly available, along with the full cohort, in a separate GEO accession upon completion of the full study from which they originate.

The scripts used for generating these results will be made publicly available via GitHub upon publication.

---

**Supplementary Figure 1.**
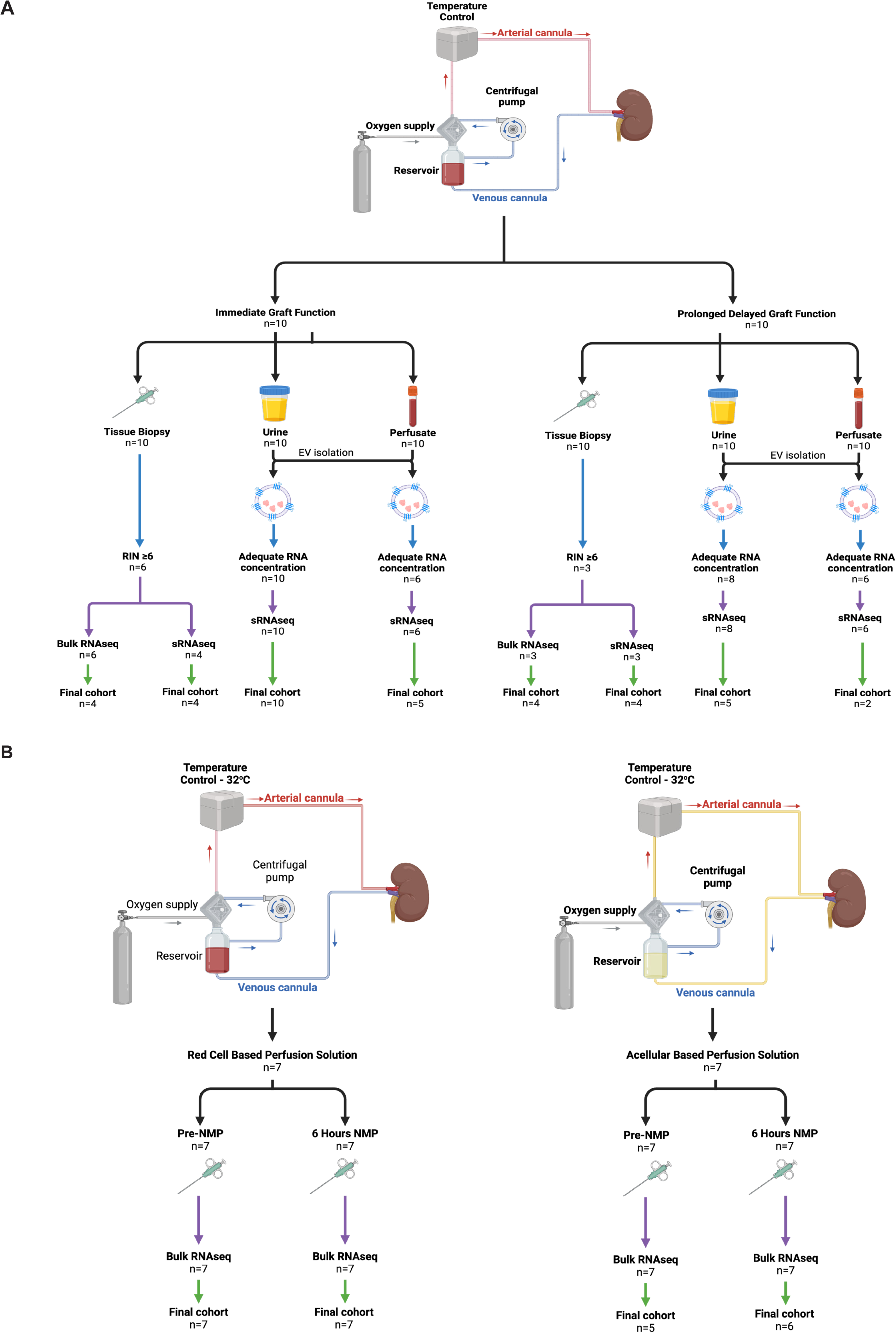
Kidney machine perfusion and sampling regime. A: Schematic of normothermic machine perfusion of human kidneys and associated sampling regime. B: Schematic of human kidney machine perfusion and sampling regime for red blood cell and acellular solution perfused kidneys. In both plots, blue arrows represent RNA extraction and quantification step, purple arrows represent submission for sequencing, green arrows represent post-sequencing technical quality control. Created in https://BioRender.com

**Supplementary Figure 2.**
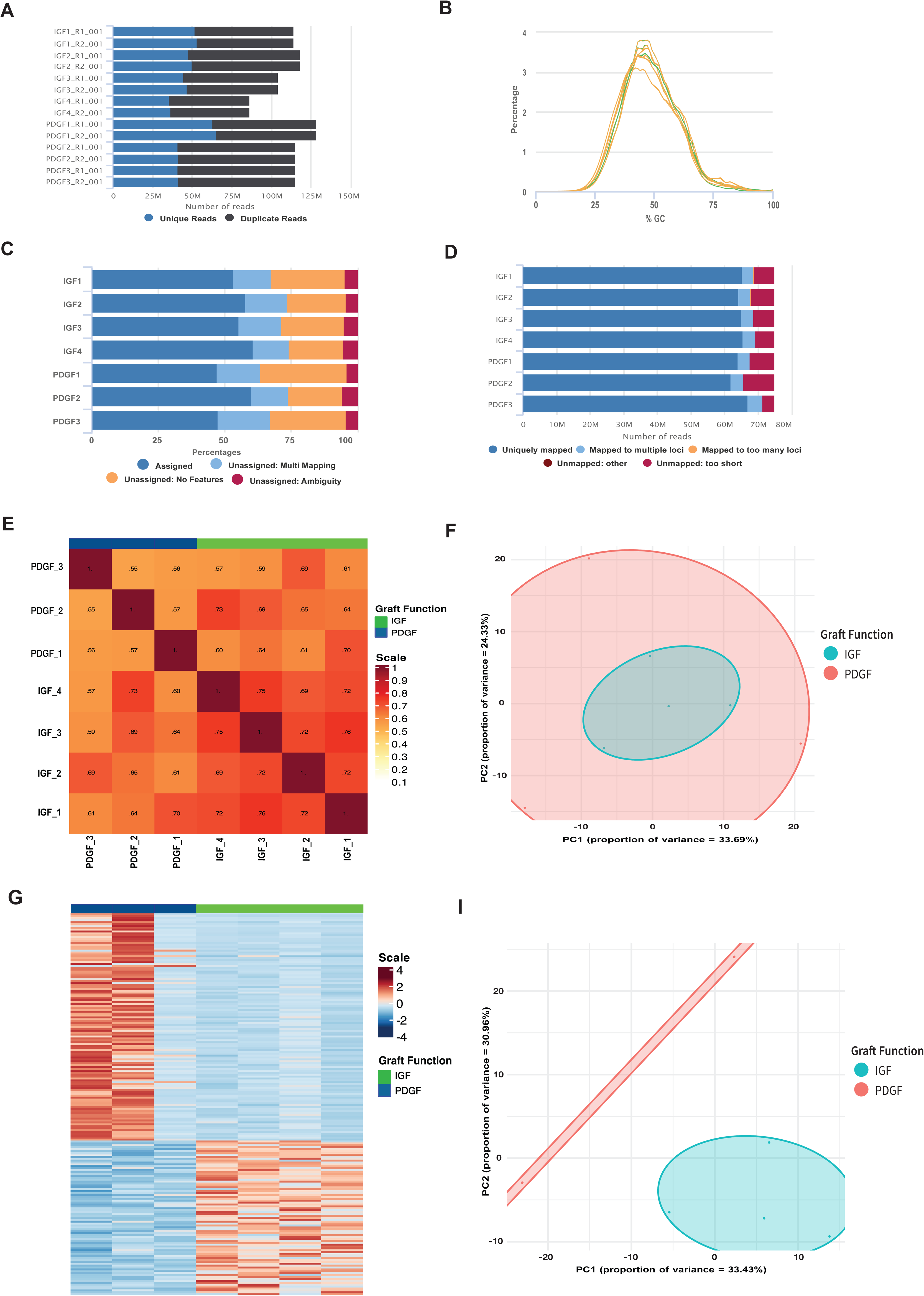
Quality control and initial analysis of tissue bulk mRNAseq data. FastQC was used for quality control of bulk mRNA sequencing outputs including sequence counts (**A**) and GC content (**B**). MultiQC aggregation of assignment (**C**) and alignment (**D**) of reads to the *H Sapiens* reference genome. Assessment of gene expression between samples by Jaccard similarity index (**E**) and principal component analysis (PCA; **F**) using top 500 most variable genes. **G**: Heatmap representation of top 200 differentially expressed genes between initial cohort of IGF and PDGF samples. **I**: PCA of final bulk mRNAseq cohort.

**Supplementary Figure 3.**
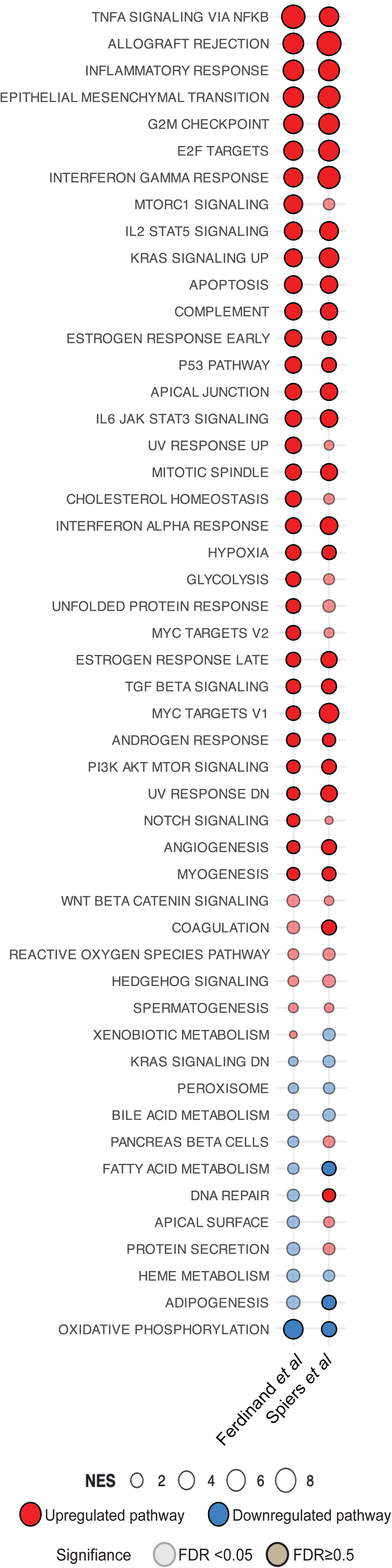
Validation of global transcriptomic signature in prolonged delayed graft function kidneys during normothermic machine perfusion. **A**: Comparison of GSEA against the Hallmarks pathways using differentially expressed genes in PDGF kidneys. Ferdinand et al data plotted from associated supplementary data(10). Size of bubble corresponds to degree of enrichment. NES, normalized enrichment score; FDR, false discovery rate. Solid colour represents FDR <0.05, transparency represents FDR ≥0.05. Full data available in Supplementary Table 3.

**Supplementary Figure 4.**
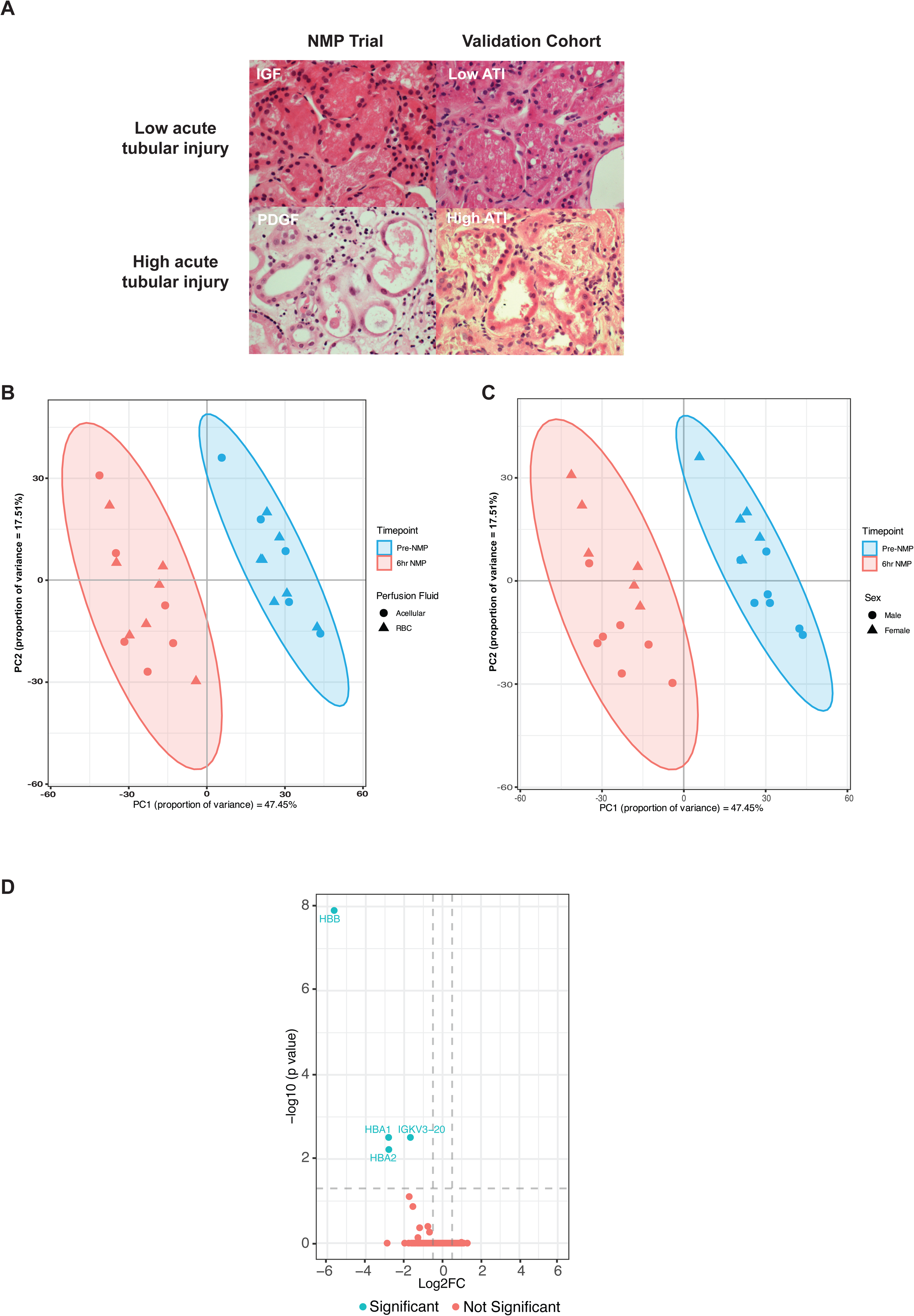
External validation cohort of machine perfused kidneys. **A**: Histological images of kidney tissue with hematoxylin & eosin stain. Top panels show representative biopsies from kidneys with low acute tubular injury (ATI) and IGF; lower panels show representative biopsies from kidneys with high ATI and PDGF. IGF, immediate graft function; PDGF, prolonged delayed graft function. Magnification x40. **B**: Principal component analysis (PCA) of top 500 genes by expression on perfusion timepoints (i.e. pre-NMP and at the end of 6hrs-NMP), annotated for perfusate. **C**: PCA of top 500 genes by expression on perfusion timepoints (i.e. pre-NMP and at the end of 6hrs-NMP), annotated for donor sex. **D**: Volcano plot of differentially expressed genes between RBC and acellular perfused kidneys in the validation cohort.

**Supplementary Figure 5.**
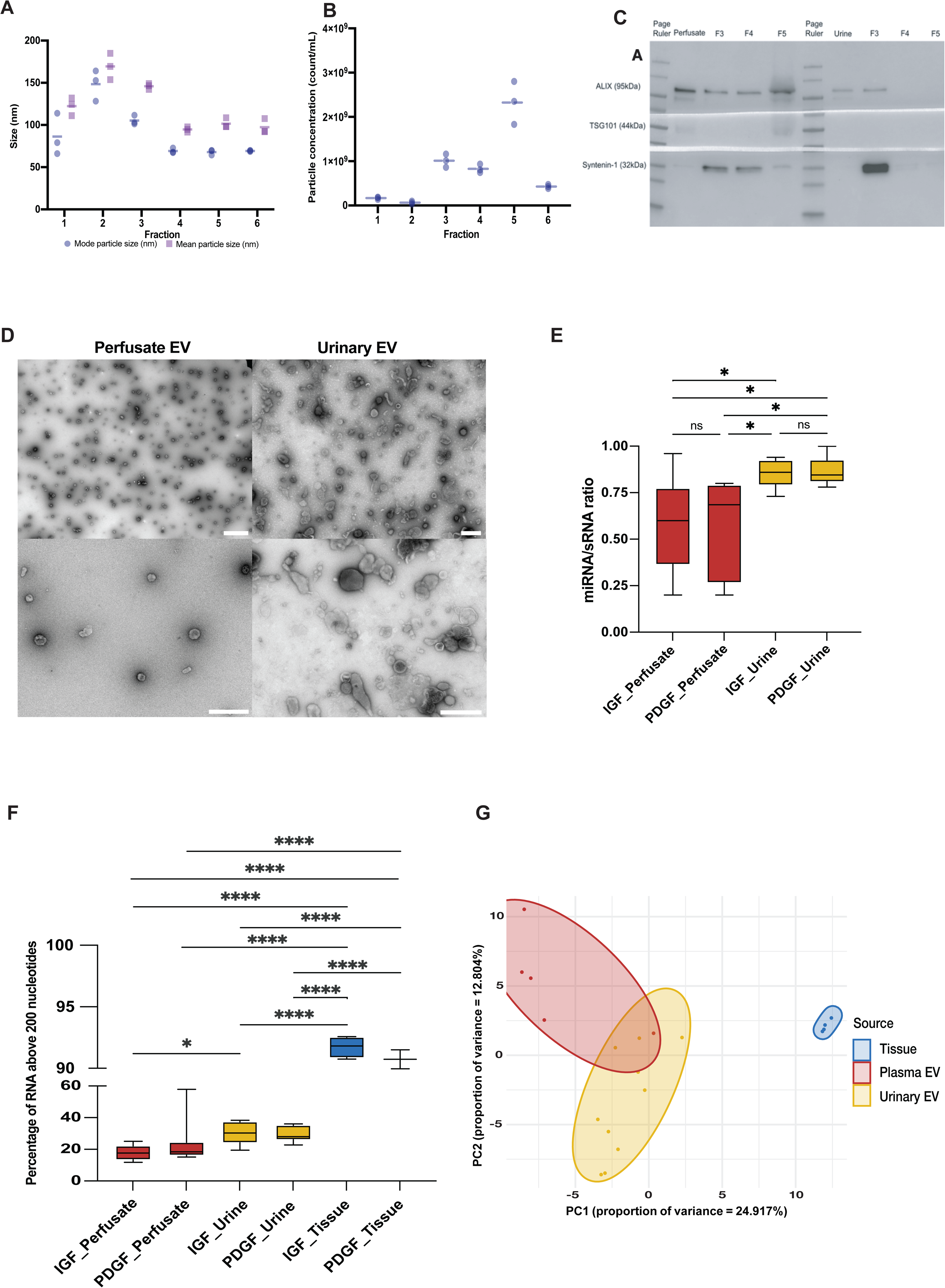
Isolation and characterisation of extracellular vesicles from biofluids of *ex situ* perfused kidneys. Mean (**A**) and mode (**B**) size of particles in fractions from urine isolated by size exclusion chromatography (SEC) determined by nanoparticle tracking analysis; EV rich fraction 3 contains particles with mean and mode size in keeping with small EVs. **C**: Western blot confirming EV specific markers (syntenin-1 and ALIX) in fractions of perfusate and urine. F3, F4, F5 correspond to SEC fraction 3, 4 and 5 respectively, with F3 enriched for EVs and F4 and F5 having increasing amounts of free protein. **D**: Transmission electron microscopy of fraction 3 from perfusate and urine confirming EV morphology. **E**: Box plot of miRNA as a percentage of total small RNA isolated from perfusate and urine derived EVs, by graft function. * = pvalue <0.05 by ANOVA. **F**: Boxplot of percentage EV RNA above 200 nucleotides in perfusate and urine derived EVs, and in kidney tissue biopsy. * = pvalue <0.05, **** = pvalue <0.0001, as determined by ANOVA. **G**: Principal component analysis of top 200 miRNAs in kidney tissue biopsies, perfusate and urine derived EVs.

**Supplementary Figure 6.**
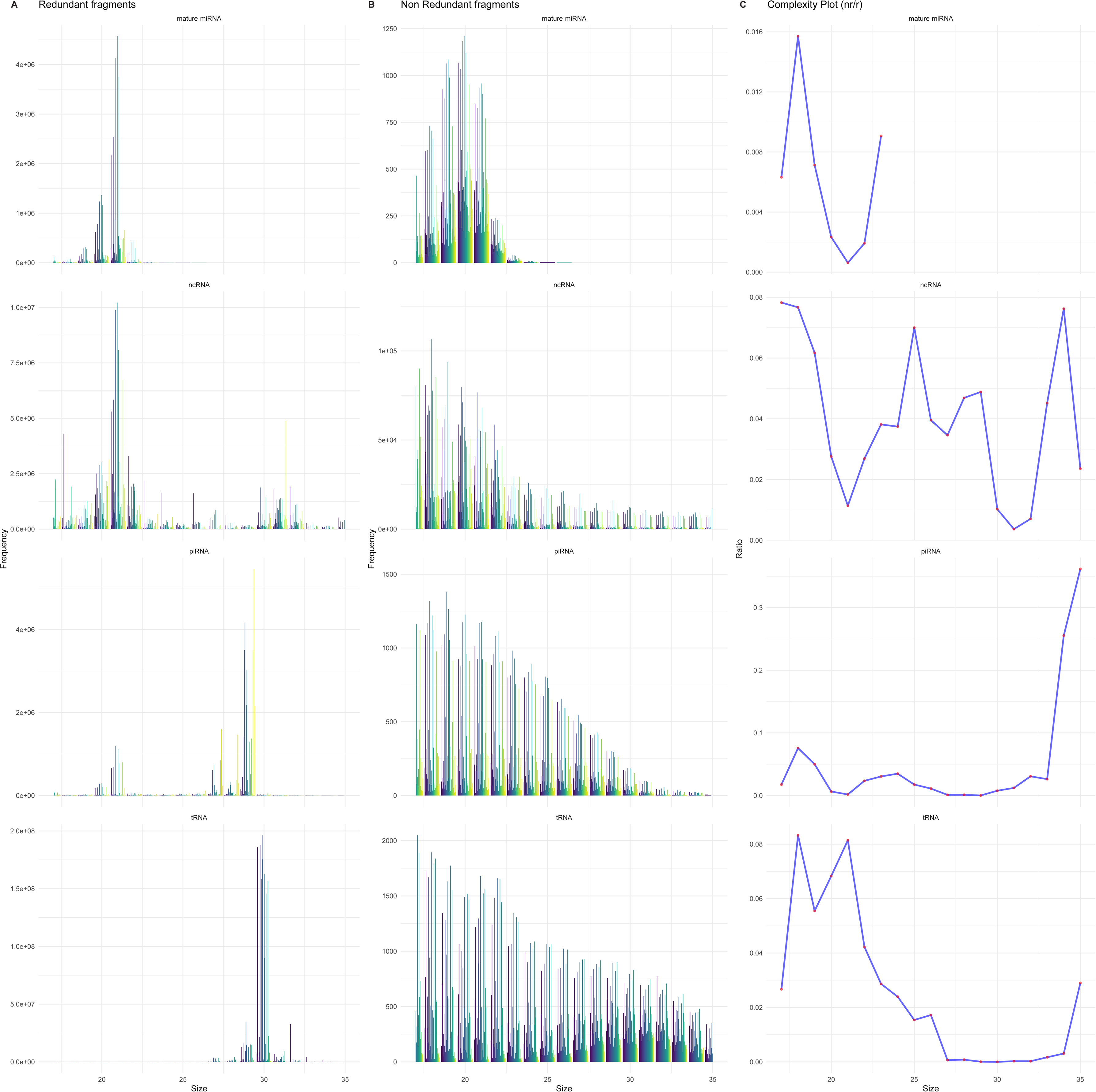
Quality control of small RNA sequencing. Redundant (**A**) and non-redundant (**B**) reads from small RNA sequencing specifically assessing mature-miRNA, non-coding (ncRNA), piRNA and tRNA species. **C**: Complexity plot demonstrating ratio of non-redundant to redundant reads for each of the small RNA species specified, by fragment size.

